# Therapeutic and Interventional Bronchoscopy Performed in Critically ill COVID-19 patients: A Systematic Review

**DOI:** 10.1101/2022.09.19.22280133

**Authors:** Saikat Samadder

## Abstract

**Background:** Coronavirus disease 2019 (COVID-19) is a highly infectious disease responsible for huge number of deaths in global population. Bronchoscopy was contraindicated for acute respiratory failure in critical patients due to possible transmission of virus to healthcare provider due to aerosol generating procedure (AGP). The safety, efficacy, complication rate, deaths, and transmission rate of virus to healthcare workers due to therapeutic and interventional bronchoscopy performed on COVID-19 patients are accessed.

**Methods:** A systematic review of literature was performed as per PRISMA 2020 guidelines. To obtain literatures available in PubMed, MEDLINE, and Google Scholars with timeline from 1st Jan 2020 – 10th Dec 2021. Databases were searched with MeSH terms bronchoscopy and COVID-19 it fetched 7350 articles. Applying primary inclusion criteria of bronchoscopy in COVID-19 patients. Secondary inclusion criteria therapeutic and interventional bronchoscopy excluding the articles on diagnostic bronchoscopy.

**Result:** Total 72 clinically relevant literatures were identified and included for further review. 1887/2558 patients underwent bronchoscopy for treatment of severe or critical COVID-19 pneumonia. therapeutic bronchoscopy was performed in 1241/1887 (65.8%) patients and interventional bronchoscopy was performed in 831/1887 (44.03%) patients. Overall, complications observed in 200/1887 (10.5%) patients. Total, 579/1887 (30%) patients died as per the literatures. Total 15 HCW (8%) were found infected during the studies. It led to successful completion of procedures in 924/940 (98.3%) patients. All three types of bronchoscopes were found to be safe for the patients. The safety, efficacy, complication rate to be 97.5%, and 98.3%, and 2.5% respectively in severely SARS-CoV-2 infected patients undergoing bronchoscopy.

**Conclusion:** This study suggests that bronchoscopy is a safe and effective procedure to be performed in patients suffering from COVID-19 pneumonia. Proper use of personal protective equipments (PPE) during bronchoscopic procedure reduced the risk of transmission of the virus from the patients to the healthcare provider.

## Introduction

In the past two years COVID-19 disease has imposed a major threat to the global population. As per WHO till 21st December 2021 there was more than 274 million infected cases and 5.3 million deaths globally. More than 8 billion doses of vaccines against SARS-CoV-2 were administered throughout the world [1]. Patients with comorbidities like diabetes, cardiovascular disease, cancer, pre-existing lung disease, liver and kidney diseases are at high risk of getting admitted to Intensive Care Unit (ICU) post acquiring severe pneumonia infection [2]. Several studies observed that aged male patients infected by SARS-CoV-2 are more likely to die and requires prolonged ICU stay regardless of treatment provided for saving these patients [3]. This virus successfully mutated as new variants known as alpha, beta, gamma, delta, epsilon, eta, theta, iota, kappa, lambda, and mu [4]. Delta variant and Omicron were reported to be more infectious than its predecessor evolving from Wuhan, China [4, 5].

Bronchoscopy is a widely used technique to diagnose different types of diseases associated with respiratory system. Bronchoscopy is one of the oldest medical device used by the surgeons since 1897 [5]. It is being successfully used to treat and diagnose various types of viral pneumonia caused by *cytomegalovirus, rhinovirus, Influenza* virus [7]. Bronchoscopes are used to diagnose tuberculosis, different types of suspected pulmonary carcinoma by bronchial wash, endobronchial ultrasound (EBUS), radial probe (RP-EBUS), transbronchial biopsy, bronchial brushing, needle aspiration, and forceps grasping of tissue for acquiring samples [8]. It is already in use for identifying the underlying diseased state of immunocompromised patients [9]. Bronchoscopy is considered AGP along with it various other procedures that are considered AGP are as follows; endotracheal intubation, open suctioning, administration of nebulized treatment, manual ventilation before intubation, turning the patient to the prone position, disconnecting the patient from the ventilator, non-invasive positive-pressure ventilation, tracheostomy, cardiopulmonary resuscitation, endo-tracheal aspiration (ETA), bronchoalveolar lavage (BAL), and bronchial aspiration (BA) [10]. Additionally, inadequate sedation, coughing during laryngoscopy, direct laryngoscopy, manual ventilation is associated risk of transmission of virus due to AGP (11). Fever and cough are the initial symptoms of this disease, presence of thick copious secretions is a major complication observed in critically-ill COVID-19 patients resulting in atelectasis.

Therapeutic and interventional bronchoscopy is an effective procedure utilized in severe or critically ill ICU admitted patients [12-15]. The use of therapeutic bronchoscopy was mainly used for the indications like acute lung failure, atelectasis, hyperaemia, routine clearance of obstructive airway in patient with lung transplantation due to COVID-19 pneumonia [16-21]. Studies suggested that the use of fibreoptic bronchoscopy helped to reduce tracheal intubation induced coughing and subsequent spread of virus [16]. In COVID-19 pneumonia patients therapeutic usage of bronchoscopy included, mucus plug and hematic secretion aspiration provided therapeutic benefit for the patients [17, 22, 23-24]. In COVID-19 patients the ETT jammed because of excessive secretions more frequently found in patients compared to pre-COVID respiratory illness [25]. While interventional procedures like reduction in lung volume, hemostasis of active bleeding, intubation, extubation, tracheostomy, FBA, surfactant delivery, and decannulation provided positive outcome for the ICU patients under prolonged mechanical ventilation [13, 26]. Percutaneous tracheostomy (PT) in the ICU patients is widely used to facilitate weaning from mechanical ventilation, to anticipate prolonged mechanical ventilation, to aid in the management of respiratory secretions and protect the airway in patients at risk of aspiration, to prevent laryngeal injury, and to minimize sedatives [27]. The therapeutic bronchoscopic procedure lasted for maximum 10 minutes, while guided PDT lasted for around 25-30 minutes for most of the COVID-19 patients [28-30].

A survey on response of physicians for use of bronchoscopy on intubated patients supported the use, found no associated risk on healthcare workers [31]. In COVID-19 pneumonia patient bronchoscopy is used for diagnosis of secondary bacterial or fungal infection, the use of diagnostic bronchoscopy was under-utilized, due to busy hospital environment and risk of AGP during the COVID-19 pandemic [32]. There are sufficient data pertaining to bronchoscopic diagnosis of secondary infections in COVID-19 patients and its efficiency to diagnose co-infection [33-34]. At present there is no review article available to determine the safety, efficacy, complications, deaths associated with the use of therapeutic and interventional bronchoscopy in COVID-19 patients. Among COVID-19 patients, gender, pregnancy, and age specific trends of death or complication post bronchoscopy is unknown. Various bronchoscopes disposable, rigid or standard utilized more frequently in COVID-19 patients was not evaluated and associated complications and risk of transmission to HCW are not assessed yet.

## Methods

Systematic review of literature was done in PubMed, MEDLINE, and Google Scholars database further in INSPERO register from 1st Jan 2020 till 10th Dec 2021 as per PRISMA 2020 guidelines [35]. In Google Scholars database searches were performed with each terms “Bronchoscopic washings”, “Bronchial aspirations”, “Therapeutic bronchoscopy”, “Toilet bronchoscopy”, “Interventional bronchoscopy”, “Foreign Body Aspiration” and “COVID-19”. In PubMed and PMC databases advanced search was performed to retrieve articles specific to bronchoscopy in COVID-19 patients with MEDLINE and time as filter (1st Jan 2020-10th Dec 2021). The specific search strings with MeSH terms (((Bronchial aspirations) OR (Pregnancy) OR (Airway management) OR (Suction) OR (Tracheostomy) OR (Trach) OR (Endotracheal tube) OR (Toilet Bronchoscopy) OR (Foreign Body) OR (Intubation) OR (Interventi*) OR (Therapeutic) AND (Bronchoscop*) AND (COVID-19) was used. During the initial screening articles in foreign language, duplicates and, in-vivo studies excluded. Under primary inclusion criteria clinical study reports presenting bronchoscopy in COVID-19 patients without any bar on age and gender included, non-bronchoscopy reports excluded. Articles suggesting practice, current understanding, guidelines, recommendations, considerations, review articles, ethical and consensus statements excluded. Secondary inclusion criteria included studies confirming COVID-19 by RT-PCR are included, studies with main focus on therapeutic and interventional bronchoscopy, excluding the articles on bronchoscopy in COVID-19 patients with diagnostic and evaluation purposes along with studies on tracheostomy. Bronchoscopy performed in patients to confirm certain suspected condition like mucosal ulceration, tracheomalacia, bronchiectasis, tracheal stenosis, pneumomediastinum etc. are excluded. Several studies isolated BAL using bronchoscope to identify microbes and modify the anti-microbial therapy for patients, such studies considered diagnostic rather than therapeutic usage. However, studies with information on both diagnostic and therapeutic/interventional bronchoscopy are included without any assumption on numbers. Articles with information on safety and efficacy were segregated and cumulative analysis of number with COVID-19 patients undergoing bronchoscopy is analyzed. Limited complications in patients and transmission of virus to healthcare professionals considered as safety information, the success rate in patients contributed to efficacy assessment cumulatively calculation shown (**Refer Supplementary Table 1-3**). The overall subjects in each studies are tabulated assessed and presented with percentage as outcome of study (**Refer figure 1**). No automation tool was used to search the literatures. Due to study type no ethical committee approval was acquired for this review. The author solely contributed all the works related to research.

**Figure 1:**
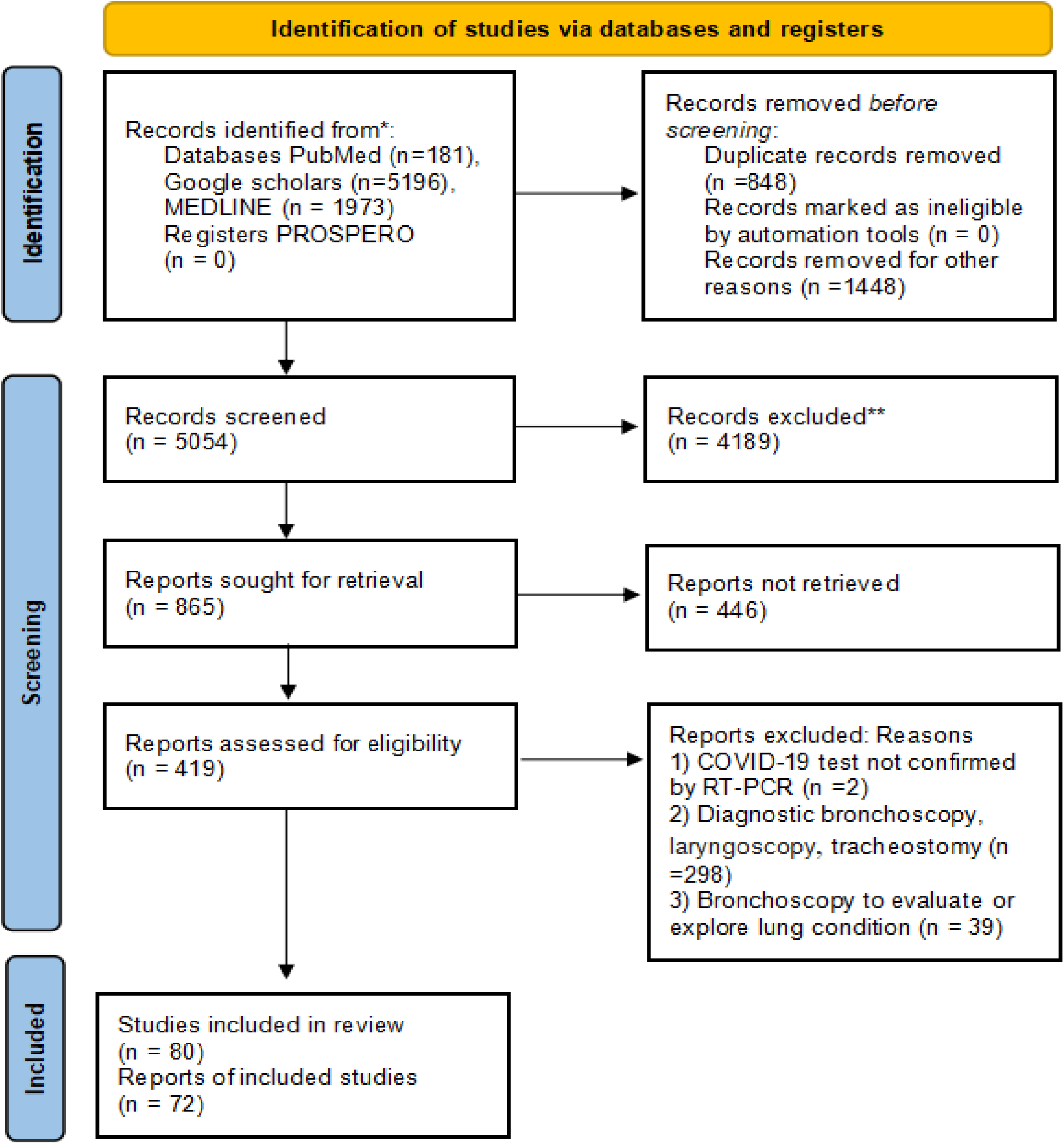
flow-chart for systematic review of literature.

### Search Results

Upon performing searches in Google Scholars, PubMed and MEDLINE found 5196, 181, and 1973 articles respectively. No systematic review found in PROSPERO register searched with terms bronchoscopy and COVID-19. 2296 articles excluded duplicates, non-English articles, and in-vivo studies. 5054 articles screened for the clinical information of bronchoscopy in COVID-19 patients. Remaining 4189 articles excluded among those review articles. Among those 865 articles sorted for retrieval of non-clinical, and other clinical studies non-specific to bronchoscopy excluded (n=446). Potentially eligible 419 articles included for detailed screening therefore downloaded for further review, under secondary exclusion criteria 336 articles excluded for not confirming COVID-19 by RT-PCR, studies on laryngoscopy, tracheostomy, diagnostics and exploratory bronchoscopy. Lastly, 80 articles included in study from search among those 72 clinical reports met all the criteria, 42 observational, prospective, retrospective, interventional, and cohort studies, 1 study was proof of concept, 8 case reports, and 21 case series found to have met the inclusion criteria are shown in **supplementary tables 4**. The data on diagnostic outcomes met rejection, only clinical information on therapeutic and interventional roles from those studies considered. The case series and case reports containing critical complications important for this study included. The flow-chart for systematic review of literature as per PRISMA 2020 guideline is presented in **Figure 1**.

## Results

72 clinical studies comprised of 2558 patients among them 1887 critically ill COVID-19 patients (74%) received various pulmonary treatment using bronchoscope. Bronchoscopy was performed in all age groups including infants, adolescents, adults, elderly male and female patients. Therapeutic bronchoscopy was performed in 1241/1887 (65.8%) patients, bronchoscopic removal of various forms of thick secretions, mucus plug, and aspiration of hematic clots performed in 1230/1241 (99.11%) patients, surfactant therapy was delivered to 7/1221 (0.56%) patients and FBA in 4 patients (0.32%) **refer Supplementary table 1 and 3**. Interventional bronchoscopy was performed in 831/1887 (44.03%) patients. Various procedures PDT 563/831 (67.7%) patients, other therapeutic interventions included ETT placement, lung volume reduction, air leakage verification, decannulation, extubation, intubation, stent/valves placement and for treatment of bleeding and airway injury in COVID-19 (28.5%) patients treated using bronchoscopy **refer Supplementary table 2 and 3**. There were 14 studies reported the use of both therapeutic and interventional bronchoscopy delivered in 871 patients, one study did not differentiate IB and TB with 31 patients **refer Supplementary table 3**.

Overall, complications found to be 200/1887 (10.5%) patients. Whereas, bronchoscopy related complications noted in 47/1837 (2.49%) patients for mild desaturation 22 patients, mild hemoptysis in 6, ETT displaced in 3, fever in 2, required intubation for 2 patient, no improvement in 4 patients, 1 bronchoscope technical problem and 7 others events related. Major bleeding, infection, and pneumothorax not observed in patients with SARS-CoV-2 infection undergoing bronchoscopy. In 11 studies involving 396 patients not reported about deaths, 32 authors reported that no deaths observed among 227 patients. Total, 579/2558 (20%) patients died as per the literatures, bronchoscopy related deaths not found. The procedure was well tolerated in most of the patients there was no death directly caused by its use of bronchoscopy. In 11 studies, 70 COVID-19 patients underwent bronchoscopy and no complications or deaths were noted (**Refer Supplementary table 1, 2 & 3**).

In 12 articles the bronchoscope was found to be both safe and effective in patients. In 26 studies the use of bronchoscope was found to be safe for 1151 patients with severe COVID-19 among them 20 authors reported in their study bronchoscopes are safe for HCWs. In 58 studies the use of bronchoscopy was successful, led to effective completion of attempted procedures in 924/940 (98.2%) patients. In 7 studies bronchoscopy showed lack of efficacy on 13 patients and 1 bronchoscopy technical problem during ongoing procedure resulted in 14/940 (1.7%) inefficacy in infected patients.

Male to female ratio was high in majority of studies, 8 studies with 397/2558 patients did not report gender of patients. In 64 studies total male 1563/2161 (72.3%) patients required more ICU admission than females therefore it is eminent that number of bronchoscopies performed more compared to female 598/2161 (27.6%) patients (**Refer Supplementary table 4**). In 33 studies 15/423 (3.5%) HCW infected with SARS-CoV-2 during their study period. Various studies referred either of the terms fibreoptic, bedside, flexible or portable bronchoscopes to designate the use of standard bronchoscopy. There is no major difference in complication rate caused by disposable bronchoscope in 4/38 (10.5%) patients reported in 17 studies, compared to standard bronchoscopes in 32/128 (19.7%) patients reported in 21 studies. Disposable bronchoscopes more frequently used in high income countries than in low or middle income nations. Majority of the studies conducted in countries like USA, Italy, and China (**refer Supplementary figure 4)**. The overall results including success rate, mortality, efficacy, complications, rate of transmission to HCWs, type of bronchoscopes used, and number of studies per country are shown in **table 1**.

**Table 1:** Overall major findings of this review are shown in above table with various study parameters such as number of patients gender, success rate, safety, mortality, complications, transmission rate, studies per country, and types of bronchoscopes as per number of studies and rate or trend.

| Overall Study findings | Number of studies | Rate (%) or overall trend |
| --- | --- | --- |
| Total number of COVID-19 patients | 72 | 2558 |
| Total number of patients undergoing bronchoscopy |  | 1887 |
| Total number of male<br>Female | 64/72 | 72 (1563 Males)<br>28 (598 Females) |
| Efficacy | 58/72 | 98.3 |
| Failure in Successful procedure |  | 1.7 |
| Safety | 26/72 | 97.5 |
| Total Mortality in COVID-19 patients | 72 | 22 |
| Mortality caused by bronchoscope | 29/72 | 0 |
| Total complications in COVID-19 patients | 33/72 | 10.5 |
| Complications caused by bronchoscope | 35/72 | 2.5 |
| Rate of transmission to HCWs performed bronchoscopy | 33/72 | 3.5 |
| Transmission caused by bronchoscopy | 33/72 | 0 |
| Number of studies/country | 72/72 | USA 24, Italy 10, China 9, Spain 4, France 1, UK 3, Ireland 1, Greece 1, Portugal 1, Belgium 3, Japan 2, South Korea 1, India 4, Indonesia 4, Pakistan 2, South Africa 1, Australia 1. |
| Types of bronchoscopes |  |  |
| Standard/flexible/fibreoptic/bedside/video-bronchoscope/portable bronchoscope | 52/72 | Middle and High income nations |
| Disposable bronchoscope | 18/72 | Upper middle and high income countries |
| Rigid bronchoscope | 4/72 | Low and High income nations. |

## Discussion

### Therapeutic Bronchoscopy in Critical patients

Therapeutic bronchoscopy is a potential procedure used to clear the airway obstruction caused by thick mucus secretions. It is useful for patients with severe or critical condition, deposition of excessive mucus or secretions are commonly seen in these COVID-19 patients (25, 18, 20-21, 36-37). Bronchoscopy should not be performed in COVID-19 patients with severe respiratory insufficiency managed under non-invasive mechanical ventilation (12). Removal of mucus plug, gelatinous mucus, thick hematic secretions caused due to hyperaemia, bronchoscopy was a lifesaving procedure for these patients [38, 39, 40, 41, 42]. Thick mucus plug resulted in airway obstruction and difficulty in ventilation (43-44). Atelectasis in patients require immediate medical attention for clearance of debris and it substantially increased oxygenation (45-47 & 17). Due to prolonged ventilation, veno venous extracorporeal mechanical oxygenation (VV-ECMO), and tracheostomy patients requires flushing out of pneumothorax (48-50). Several studies reported usefulness of therapeutic bronchoscopy among those very few found bronchoscopy ineffective and technical problem in patients [51, 11, 21, 25]. Bhatraju et al. reported in their study 17 patients required bronchoscopy without any secondary infection, however patients required vasopressors. Active use of heated humidification or moist heated humidifiers was used in several studies that reduced bronchoscopy time (23, 25). Bronchoscopy was successfully performed in 2 positive patients for mucus secretions aspiration and FBA removal (52). Foreign body aspiration was done only in few patients with COVID-19, mostly FBA was done once the RT-PCR test turned negative [53]. The presence of haematic secretions in the distal bronchial tract was found to the reason of high mortality in patients (50). In one COVID-19 patient chest drain was performed due to pneumothorax but oxygenation status did not change post bronchoscopic suction of mucus plug (54). Pulmonary toilet was done by the use of bronchoscopes to remove mucus and secretions in COVID-19 patients [55, 29-30].

In a study laryngoscopy and tracheostomy resulted in tissue laceration resulting in death of 4 patients, the patients who died are older than patients who survived (56). There was no significant difference in-hospital mortality or ICU admission of COVID-19 positive patients compared to negative patients respectively [50]. The COVID-19 disease trends to affect less and has mild clinical course in pregnant women compared to influenza [57]. Post-partum COVID-19 related complications exacerbates, severity in these women are rare during pregnancy and therapeutic bronchoscopy could be performed successfully post-delivery [45-46, 58-59]. In infants or pediatric patients bronchoscopy proved to be a safe therapeutic device used for mucus plug removal [40, 52-53, 60]. In elderly patients with COVID-19 requiring lung transplant underwent routine therapeutic bronchoscopy no complications seen in patients caused by its regular usage (19-20).

### Interventional Bronchoscopy in Critical Patients

Interventional bronchoscopy is a useful technique to reduce the disease burden of severe or critical SARS-CoV-2 pneumonia [61, 13]. Bronchoscopy guided tracheostomy is a very useful technique for reducing mortality in ICU [62-63, 64, 11]. Various types of interventional bronchoscopy performed in critical COVID-19 patients to complement endotracheal intubation (ETT), orotracheal intubation, (OTT) repositioning, bronchoscopic percutaneous dilational tracheostomy, stent and valve placement, embolization of airway bleeding, air leakage detection, lung volume reduction, bronchoscopic assessment pre-decannulation and decannulation were performed [25, 12, 50, 65-66, 56, 22]. Meyer et al. reported bronchoscope guided PDT decreased the risk of bleeding in comparison to open tracheostomies, considering the high rate of anti-coagulation therapy required for these patients. Guided PDT reduced transmission of virus to HCWs from critically ill COVID-19 patients [15, 68-69], although there was no major difference in complication rate in PDT compared to open tracheostomy [64, 61]. However, in a pre-COVID study demonstrated that use of bronchoscopes did not affect the overall outcome in patients with open or percutaneous tracheostomy [70]. Early PDT for COVID-19 patients was found related to reduced stay in ICU than those underwent delayed PDT [71]. The use of bronchoscopes for ETT placement reduced aerosolization process (69, 68). Therefore, possibly no transmission to bronchoscopists could be identified in studies related to bronchoscopy guided PDT in infected patients (**Refer Supplementary table 2-3**). The survival rate post-tracheobronchial stents placement found to be only 3-4 months in patients with malignant airway disease; the one-year survival rate was reported to be 15% (65). In few studies mortality rate post-tracheostomy was reported to be below 8% in COVID-19 patients (72).

### Complications Observed in Patients

In a randomized controlled study with COVID-19 and non-COVID-19 patients found tracheal and thoracic complications only in COVID-19 patients (73). Tracheal complications included lesions and tracheoesophageal fistulas, the thoracic complications included pneumothorax, pneumomediastinum, and subcutaneous emphysema are commonly observed in COVID-19 patients during bronchoscopy (73, 56). The COVID-19 patients supine or prone positioned during bronchoscopy did not change the course of patient’s hospital stay and no complications observed (74, 28). Bronchoscopy was not performed in pregnant women with severe COVID-19, bronchoscopy was considered safe post-partum. Therefore, no complications related to bronchoscopy could be observed during gestational period (45-46, 59). Bronchoscopy guided intubation in COVID-19 patients could be considered safe strategy, as mild desaturation could be observed [11, 75]. Therapeutic bronchoscopy delivered to pediatric or young patients was safe for severe COVID-19 pneumonia (60, 40).

The main complications observed in patients related to bronchoscopy in COVID-19 patients were no improvement in very few patients, minor bleeding, cellulitis, drop in oxygen saturation (SpO2) <90%, or desaturation, pneumothorax, site irritation, mild respiratory failure, and difficult to swallow were common during pre-COVID-19 era [71, 76, 35, 77, 21]. Several studies reported patient underwent bronchoscopic aspiration of secretions but died due to disease progression without mentioning any complication related to bronchoscope [78, 79, 26]. Technical problem and difficulty in swallowing two complications of bronchoscope observed in two studies, both used disposable bronchoscopes [25]. Complications were more often due to PDT and tracheostomy leading to major or minor bleeding, delayed complications, cellulitis, infection and pneumothorax [71, 27, 80, 69, 81-84, 73]. Events valve migration, ETT dislodgement in patients was not associated to bronchoscopy [85, 49]. Major bleeding, hemoptysis, and delayed bleeding was rare after therapeutic bronchoscopy and was found in patients undergoing bronchoscopy guided PDT (49, 77). Bronchoscopy guided tracheostomy caused majority of adverse events in COVID-19 patients compared to other interventional bronchoscopy or therapeutic bronchoscopic procedures (61, 86, 3, 82, 81, 56, 87). Bronchoscopes were not responsible for occurrence of any major events in severely ill patients undergone PDT (**Refer Supplementary figure 2-3**). Numerous articles could be found related to open and guided tracheostomy a separate meta-analysis should be conducted in COVID patients to understand the safety and efficacy of both procedures. The major complications found are not associated to its usage.

### Mortality Rate

None of the articles reported the death of the patients due to the use of bronchoscopes. Death of the patients were mainly related to disease progression in patents and due to prolonged stay in ICU [17, 45, 59, 50]. There was no mortality reported in bronchoscopists performing the maneuvers. Age, mucus plugs, hematic secretions and mucosal hyperaemia was found to be major contributor for in-hospital mortality of COVID-19 pneumonia patients (50). Bronchoscopy was not considered as a worse prognostic factor for predicting mortality in COVID-19 patients (25). Bahatraju et al. reported in their multi-center case series deaths occurred in 12 out of 24 patients with do not resuscitate orders, two of their patients received TB it is unknown if patients died, as the clinical course did not change for these patients. Botti et al. reported patients did not die due to bronchoscopy or open/closed tracheostomy and possible cause of death was severe ARDS. Deaths were reported to be caused by ARDS, increase in viral replication, multi-organ failure, mechanical ventilation, open tracheostomy and PDT (45, 88, 71, 72, 3, 83, 81). Lacerations caused deaths of patients, could be due to the infection or post tracheostomy event not relevant to therapeutic usage of bronchoscopes [56, 73].

### Infection in Healthcare Providers

Overall, 15 HCW reportedly infected with SARS-CoV-2 virus during the course of their routine participation in study. However, none of the studies reported that any of these transmission could be due to the bronchoscopic procedure [16, 24, 76, 28]. These infections were not transmitted from patients in ICU wards as there was no outbreak of viral infections observed among other HCW present associated with the same bronchoscopic procedures [52]. Thereby, suggesting that the procedure is safe for the bronchoscopists, as maximum precautionary measures were adapted to reduce the aerosolization process [26, 52-53, 55, 59, 89].

Additionally, it was well documented in each of the studies that proper use of personnel protection equipment (PPE), googles, double gloves, cap, shoe cover, visor, gown, and filtering face piece level 3 (FFP3) mask [54, 56]. Use of positive pressure hood, bronchoscopy under negative pressure ventilation, regular body temperature checks for HCWs, and weekly RT-PCR. Compliance with reprocessing of bronchoscopes reduced the rate of transmission and co-infection in patients (3, 68, 86, 75, 29). The HCWs performing high risk maneuvers during 2020-2021 pandemic were comparatively safer than those performed tracheal intubations in SARS-CoV-1 epidemic (90). During SARS-CoV-1 epidemic 53 HCWs infected from the high risk procedure such as intubation (91).

### Types of Bronchoscopes Used for Therapeutic Benefits

Disposable bronchoscopy was used in 14 studies mainly high income countries, utilized single use bronchoscopes, while middle income countries used traditional standard bronchoscopes. Rigid bronchoscopy used in small number of patients in 6 studies all patients survived without any major complication **refer supplementary table 1**. Disposable bronchoscopes were found to have least number of complications caused by it (86, 25), compared to standard bronchoscopes which were responsible for few mild-moderate events (14, 74, 21, 11, 51, 45, 76, 56). Cheap and affordable re-useable bronchoscopes would be beneficial for reducing infections lower or middle income countries, presently are less utilized. Disposable or standard fibreoptic bronchoscopes usage did show any major difference in transmission rate to HCWs.

### Limitations

Long-term follow-up data on patient’s death was not available in many articles, due to their study design therefore the mortality rate is calculated based on data available. In few studies gender of subjects was not available so it was difficult to identify exact number of males and females included in the studies. In majority of the studies number of bronchoscopists participating in the studies not reported. There could be an overall risk of bias due to low number of randomized controlled trial included in this review, inconsistency, indirectness, and imprecision not assessed for each studies included.

## Conclusion

Bronchoscopy is a very potential tool available for diagnosis and treatment of various respiratory diseases [22, 79]. It was widely used during the pandemic to diagnose co-infection in patients, to overrule false RT-PCR results from the nasopharyngeal samples and to change antibiotics to mitigate life threatening situations caused by co-infection [38]. This study is important because until December 2021 there were no anti-viral drug available in market globally to treat the critically ill COVID-19 pneumonia patients, and bronchoscopy demonstrated a reliable tool for treatment of various critical conditions. The procedure was well tolerated in most of the patients there was no major complications or death directly caused by its usage. There were no severe irreversible or new adverse event found related to bronchoscopy treatment in patients with critical COVID-19. This study favors the use of bronchoscope for the clearance of lung blockage and for rescuing oxygenation status in patients, and helped to overcome other detrimental situations resulted by interventional procedures or prolonged ICU stay and progression of viral pneumonia [12]. The use of bronchoscope reduced the lung blockage caused by various forms of increased mucus secretions these are more commonly seen only in COVID-19 pneumonia patients, unlike patients with other respiratory diseases [74, 92, 25, 17]. Therefore, treatment using heat moist exchanger or humidifiers, surfactant, expectorant, and nebulization therapy could be useful for aspiration of thick mucus reducing the time of bronchoscopic procedure and increasing overall efficacy [26, 36, 23, 25]. During the early phase of COVID-19 pandemic bronchoscopy and laryngoscopy use was reduced (32, 91). In future epidemic and pandemic situations, bronchoscope could be effectively used without delay as per the need in ICU under appropriate precautionary measures.

## Data Availability

All data produced in the present work are contained in the manuscript

## Acknowledgement

The author would like to thank Dr. Levra (University of Torino, Italy) for clarification of doubts. I extend my gratitude and appreciation to all health-care workers who provided and cared for the study subjects during the COVID-19 pandemic despite of transmission risk.

---

**Supplementary table 1:**
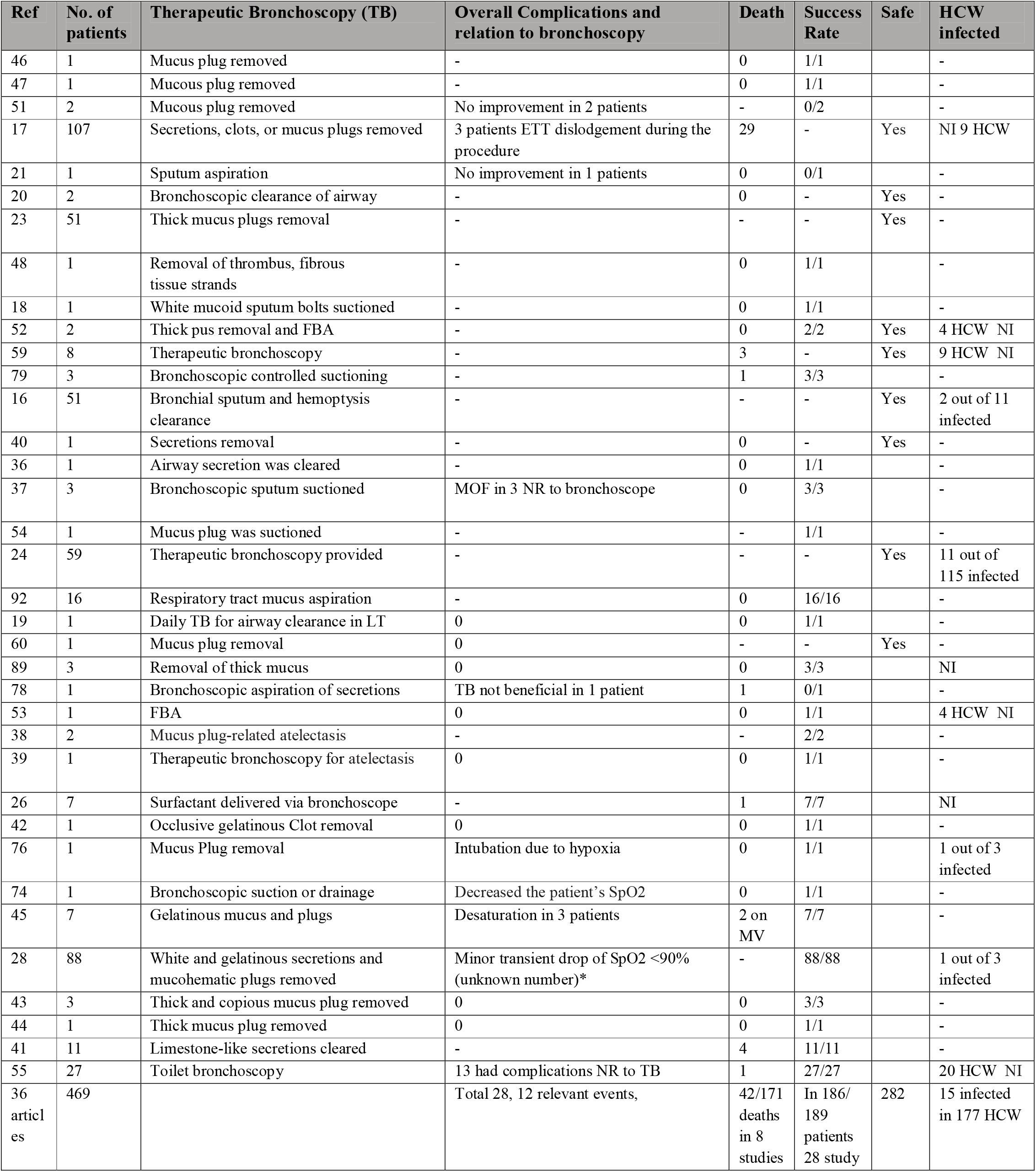
Clinical Studies with Therapeutic Bronchoscopy: Total number of patients undergoing bronchoscopy. In mortality column 0 represents no deaths observed, NI: Not Infected, HCW: Healthcare worker, NR: Not related, (-) Not reported, ETT: Endotracheal tube, SpO2: Oxygen saturation, MOF: Multi-organ failure, MV: Mechanical ventilation, FBA: Foreign body aspiration, ICU: Intensive care unit, LT: Lung transplant.

**Supplementary table 2:** Clinical Studies with Interventional Bronchoscopy: Total number of patients undergoing bronchoscopy. In mortality column 0 represents no deaths observed, PT: Percutaneous tracheostomy, EBV: Endobronchial valves, (-) Not reported, NI: Not infected, Acute respiratory distress syndrome, ARDS; MOF: Multi-organ failure. (*) complications in unknown number of patients [28].

| Ref | No. of patients | Interventional Bronchoscopy use | Overall Complication and relation to bronchoscopy | Death | Success rate | IB Safe | HCW infected |
| --- | --- | --- | --- | --- | --- | --- | --- |
| 61 | 17 | Bronchoscopy guided PDT | 9 complications due to PDT | 3 | 17/17 | - | 3 HCW NI |
| 86 | 25 | Bronchoscopy guided PDT | PDT related minor bleeding in 1 patient, desaturation in 2 patients related to IB | 9 | 25/25 | Yes | 4 HCW NI |
| 75 | 12 | Bronchoscopy guided tracheal intubation | - | - | 12/12 | Yes | 9 HCW NI |
| 3 | 29 | Bronchoscopy guided PDT | 1 bleeding 1 cellulitis NR to IB | 6 due to PDT | 29/29 | Yes | 4 HCW NI |
| 49 | 2 | Bronchoscopy guided air leak using a balloon occlusion test and valve placement | Valves migrated in 1 patient NR to IB | 0 | 2/2 | Yes | - |
| 73 | 30 | Bronchoscopy guided PDT | Late tracheal complication in 28 due to COVID-19 NR to PDT or IB | 8 | 30/30 | - | - |
| 27 | 16 | Bronchoscopic assistance in PDT | 1 patients had infection, and 2 bleeding due to PDT | 4 | 16/16 | Yes | All 4 HCW NI |
| 62 | 3 | Bronchoscopic assistance in PDT | 0 | 0 | 3/3 | Yes | All 4 HCW NI |
| 82 | 7 | Bronchoscopic assistance in PDT | Bleeding in 2 NR to bronchoscope | 5 | - | Yes | All 5 NI |
| 72 | 2 | Bronchoscopic treatment for bleeding | - | 0 | 2/2 | - | 2 HCW NI |
| 81 | 12 | Bronchoscopy guided PDT | Brief bleeding due to PDT in 1 patient | 2 due to ARDS | - | Yes | 24 HCW NI |
| 67 | 5 | PDT under bronchoscopic assistance in 1 and 4 other additional IB | - | 0 | 5/5 | - | 6 HCW NI |
| 83 | 11 | Bronchoscopy guided PDT | 2 bleeding and 1 pneumothorax NR | 3 | 11/11 | Yes | All 4 NI |
| 84 | 58 | Bronchoscopy guided PDT | Minor bleeding 12, Site irritation in 6 patients, repeat procedure in 5 NR | 0 | 58/58 | - | All 3 NI |
| 68 | 11 | Bronchoscopy guided PDT | Delayed bleeding in 1 patient NR | 0 | 11/11 | Yes | All 4 NI |
| 87 | 9 | Confirm ETT placement for tracheostomy with bronchoscope | 2 late bleeding NR to IB | 4 | 9/9 | - | All 3 NI |
| 85 | 1 | Air leakage detection and lung volume reduction with EBV | Valves migrated NR to IB | 0 | 1/1 | - | - |
| 71 | 120 | Fibro-bronchoscopy guided PDT | Minor bleeding 23 patients, subcutaneous emphysema in 3, dysphagia in 11 patients. All were PDT related. | 42 deaths COVID-19 related | - | Yes | NI 24 HCW |
| 88 | 38 | Bronchoscopic assistance in PDT | Skin bleeding in 2 NR to bronchoscopy | Intracranial hemorrhage 2, MOF 4 | 38/38 | - | 2 HCW NI |
| 63 | 3 | Bronchoscopy guided PT | - | 0 | 3/3 | - | All 3 HCW NI |
| 11 | 58 | Bronchoscopy guided intubation | SpO2 <90% in 9 patients related to IB | - | 49/58 | - | NI 6 HCW |
| 64 | 28 | Bronchoscopy guided tracheostomy | 2 patients bleeding NR to IB | 2 | 28/28 | - | NI |
| 22 articles | 497 | PDT in 409 patients + Other IBs 88 patients | 129 patients in 17 studies, 11 events from IB observed | 94/427 in 13 studies | 349/358 in 19 studies | 248 | 0/111 infected |

**Table 3:**
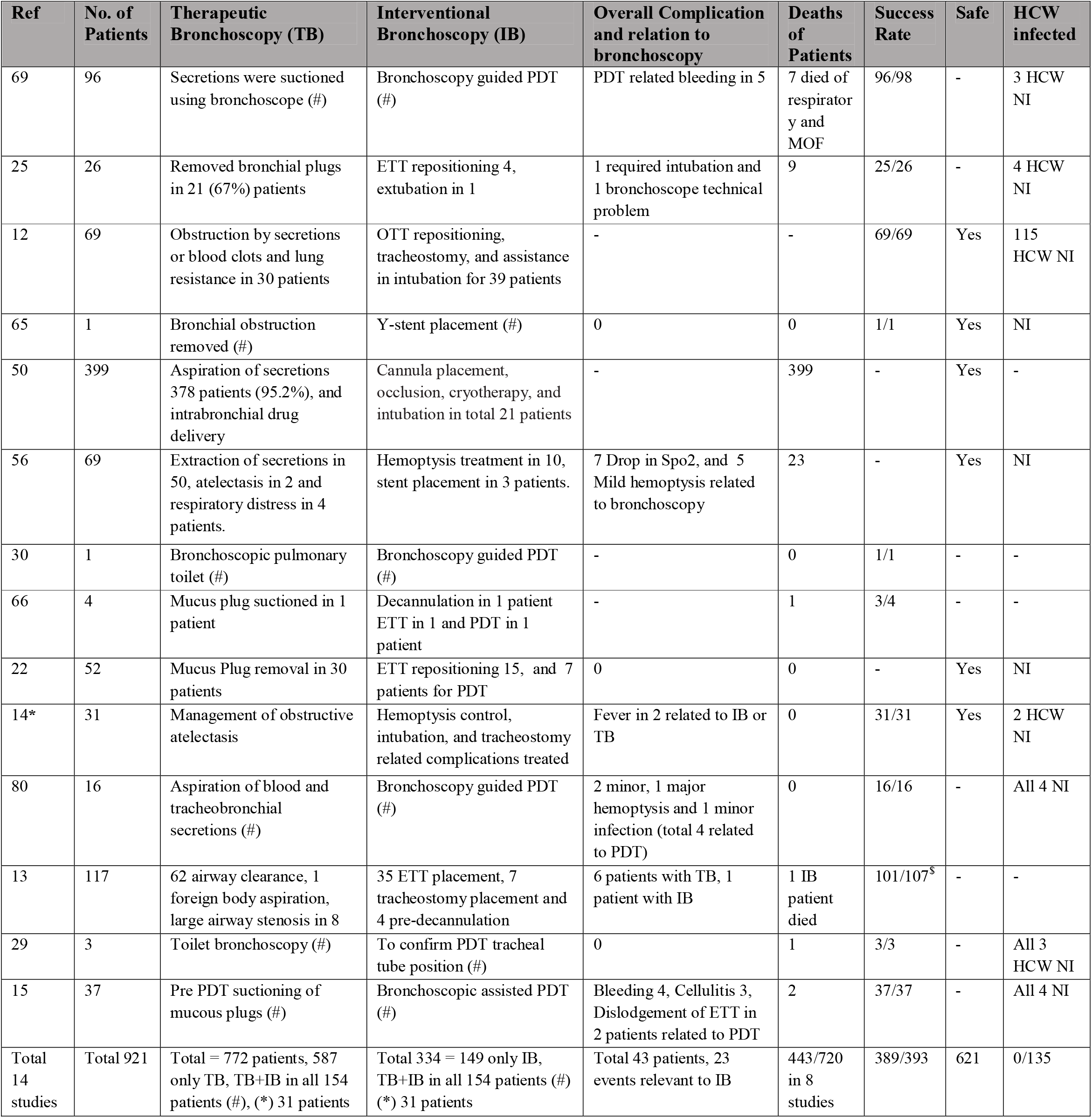
Clinical Studies with both Therapeutic and Interventional use of Bronchoscopy, Total number of patients undergoing bronchoscopy. In mortality column 0 represents no deaths observed, PDT: Percutaneous dilational tracheostomy, (**-**) Not reported, NI; represents Not Infected, (*) represents the number of patients receiving IB is unknown [14]. Post bronchoscopy clinical decline within 12 hrs ($). The studies in which both the procedures performed in patients are mentioned as (#) represents number of patients undergoing both TB and IB.

| Ref | No. of Patients | Therapeutic Bronchoscopy (TB) | Interventional Bronchoscopy (IB) | Overall Complication and relation to bronchoscopy | Deaths of Patients | Success Rate | Safe | HCW infected |
| --- | --- | --- | --- | --- | --- | --- | --- | --- |
| 69 | 96 | Secretions were suctioned using bronchoscope (#) | Bronchoscopy guided PDT (#) | PDT related bleeding in 5 | 7 died of respiratory and MOF | 96/98 | - | 3 HCW NI |
| 25 | 26 | Removed bronchial plugs in 21 (67%) patients | ETT repositioning 4, extubation in 1 | 1 required intubation and 1 bronchoscope technical problem | 9 | 25/26 | - | 4 HCW NI |
| 12 | 69 | Obstruction by secretions or blood clots and lung resistance in 30 patients | OTT repositioning, tracheostomy, and assistance in intubation for 39 patients | - | - | 69/69 | Yes | 115 HCW NI |
| 65 | 1 | Bronchial obstruction removed (#) | Y-stent placement (#) | 0 | 0 | 1/1 | Yes | NI |
| 50 | 399 | Aspiration of secretions 378 patients (95.2%), and intrabronchial drug delivery | Cannula placement, occlusion, cryotherapy, and intubation in total 21 patients | - | 399 | - | Yes | - |
| 56 | 69 | Extraction of secretions in 50, atelectasis in 2 and respiratory distress in 4 patients. | Hemoptysis treatment in 10, stent placement in 3 patients. | 7 Drop in Spo2, and 5 Mild hemoptysis related to bronchoscopy | 23 | - | Yes | NI |
| 30 | 1 | Bronchoscopic pulmonary toilet (#) | Bronchoscopy guided PDT (#) | - | 0 | 1/1 | - | - |
| 66 | 4 | Mucus plug suctioned in 1 patient | Decannulation in 1 patient ETT in 1 and PDT in 1 patient | - | 1 | 3/4 | - | - |
| 22 | 52 | Mucus Plug removal in 30 patients | ETT repositioning 15, and 7 patients for PDT | 0 | 0 | - | Yes | NI |
| 14* | 31 | Management of obstructive atelectasis | Hemoptysis control, intubation, and tracheostomy related complications treated | Fever in 2 related to IB or TB | 0 | 31/31 | Yes | 2 HCW NI |
| 80 | 16 | Aspiration of blood and tracheobronchial secretions (#) | Bronchoscopy guided PDT (#) | 2 minor, 1 major hemoptysis and 1 minor infection (total 4 related to PDT) | 0 | 16/16 | - | All 4 NI |
| 13 | 117 | 62 airway clearance, 1 foreign body aspiration, large airway stenosis in 8 | 35 ETT placement, 7 tracheostomy placement and 4 pre-decannulation | 6 patients with TB, 1 patient with IB | 1 IB patient died | 101/107<sup>\$</sup> | - | - |
| 29 | 3 | Toilet bronchoscopy (#) | To confirm PDT tracheal tube position (#) | 0 | 1 | 3/3 | - | All 3 HCW NI |
| 15 | 37 | Pre PDT suctioning of mucous plugs (#) | Bronchoscopic assisted PDT (#) | Bleeding 4, Cellulitis 3, Dislodgement of ETT in 2 patients related to PDT | 2 | 37/37 | - | All 4 NI |
| Total 14 studies | Total 921 | Total = 772 patients, 587 only TB, TB+IB in all 154 patients (#), (*) 31 patients | Total 334 = 149 only IB, TB+IB in all 154 patients (#) (*) 31 patients | Total 43 patients, 23 events relevant to IB | 443/720 in 8 studies | 389/393 | 621 | 0/135 |

**Supplementary table 4:**
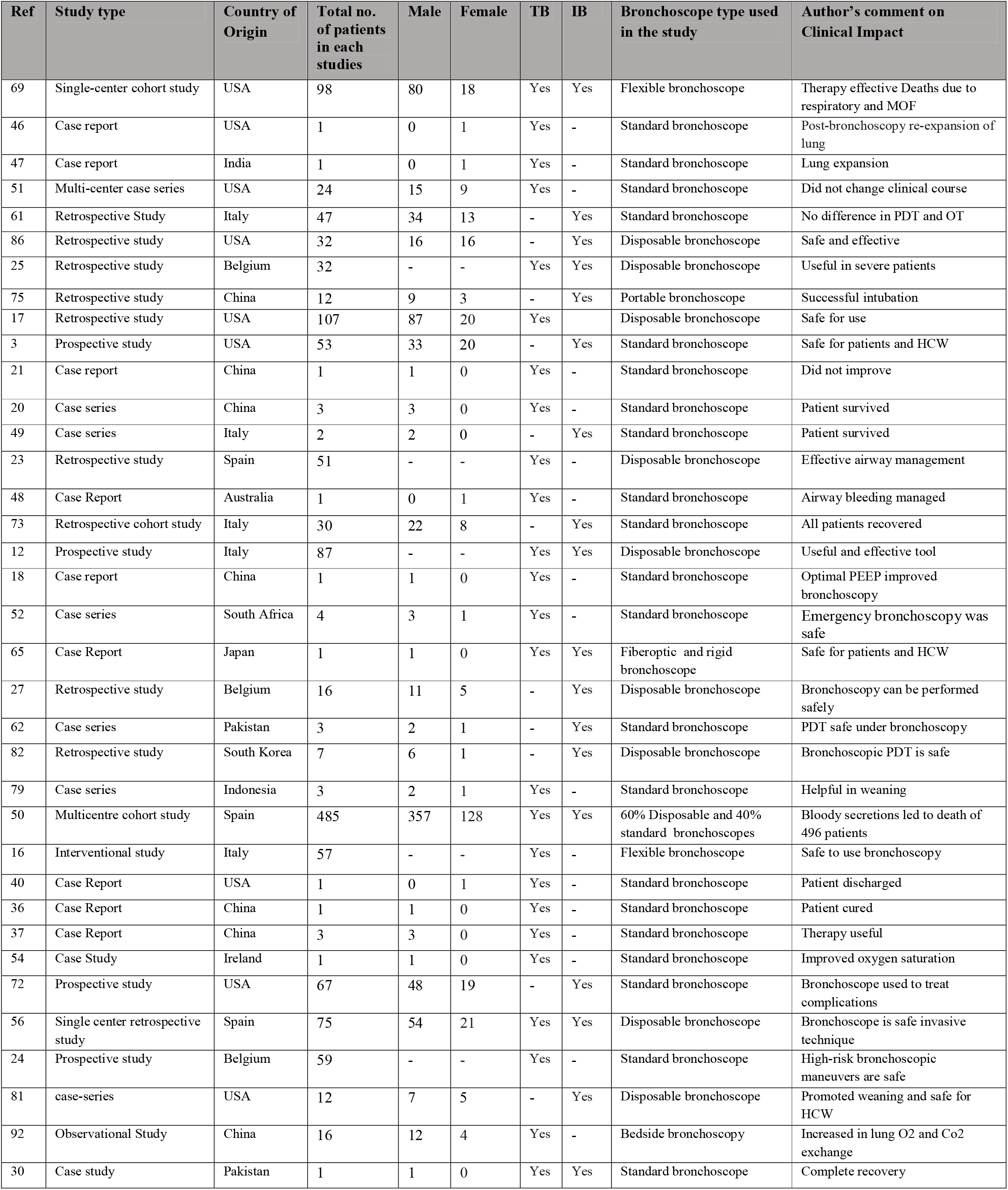
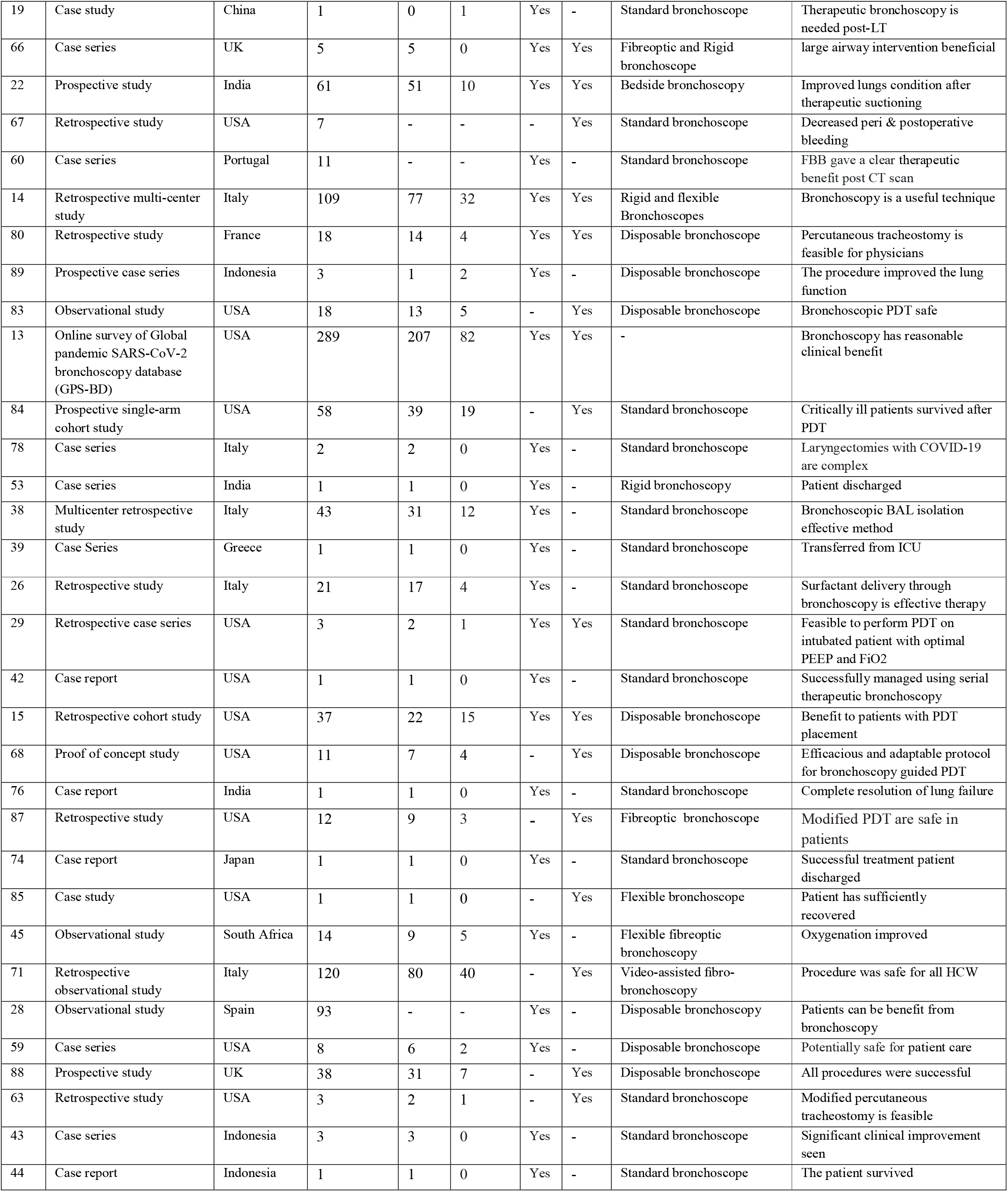
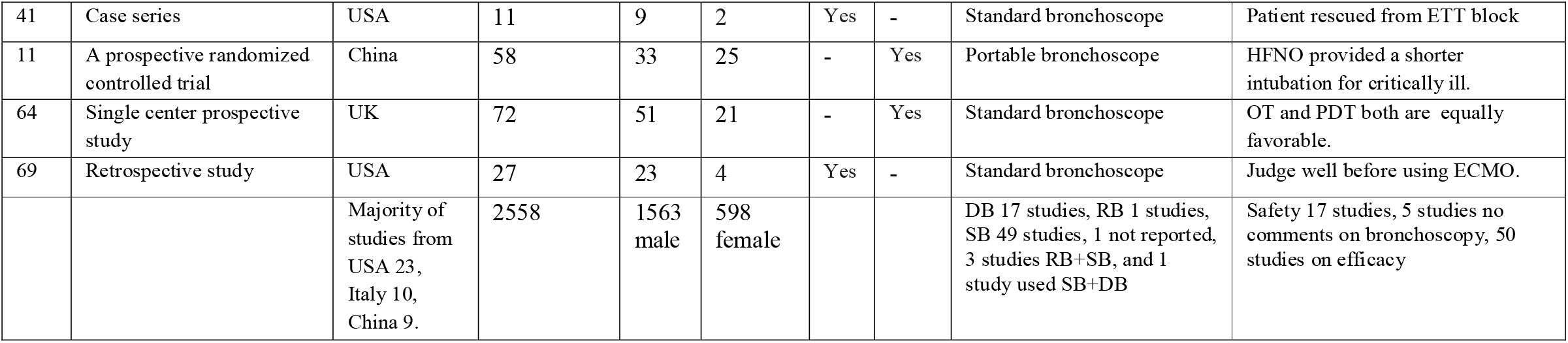
Clinical Studies with both Therapeutic and Interventional use of Bronchoscopy mentioned as “Yes”. “F” represents female and “M” represents male. (-) Not reported, In 7 articles gender not mentioned. DB: Disposable bronchoscope, SB: Standard bronchoscope, RB: Rigid bronchoscope. The

| Ref | Study type | Country of Origin | Total no. of patients in each studies | Male | Female | TB | IB | Bronchoscope type used in the study | Author's comment on Clinical Impact |
| --- | --- | --- | --- | --- | --- | --- | --- | --- | --- |
| 69 | Single-center cohort study | USA | 98 | 80 | 18 | Yes | Yes | Flexible bronchoscope | Therapy effective Deaths due to respiratory and MOF |
| 46 | Case report | USA | 1 | 0 | 1 | Yes | - | Standard bronchoscope | Post-bronchoscopy re-expansion of lung |
| 47 | Case report | India | 1 | 0 | 1 | Yes | - | Standard bronchoscope | Lung expansion |
| 51 | Multi-center case series | USA | 24 | 15 | 9 | Yes | - | Standard bronchoscope | Did not change clinical course |
| 61 | Retrospective Study | Italy | 47 | 34 | 13 | - | Yes | Standard bronchoscope | No difference in PDT and OT |
| 86 | Retrospective study | USA | 32 | 16 | 16 | - | Yes | Disposable bronchoscope | Safe and effective |
| 25 | Retrospective study | Belgium | 32 | - | - | Yes | Yes | Disposable bronchoscope | Useful in severe patients |
| 75 | Retrospective study | China | 12 | 9 | 3 | - | Yes | Portable bronchoscope | Successful intubation |
| 17 | Retrospective study | USA | 107 | 87 | 20 | Yes |  | Disposable bronchoscope | Safe for use |
| 3 | Prospective study | USA | 53 | 33 | 20 | - | Yes | Standard bronchoscope | Safe for patients and HCW |
| 21 | Case report | China | 1 | 1 | 0 | Yes | - | Standard bronchoscope | Did not improve |
| 20 | Case series | China | 3 | 3 | 0 | Yes | - | Standard bronchoscope | Patient survived |
| 49 | Case series | Italy | 2 | 2 | 0 | - | Yes | Standard bronchoscope | Patient survived |
| 23 | Retrospective study | Spain | 51 | - | - | Yes | - | Disposable bronchoscope | Effective airway management |
| 48 | Case Report | Australia | 1 | 0 | 1 | Yes | - | Standard bronchoscope | Airway bleeding managed |
| 73 | Retrospective cohort study | Italy | 30 | 22 | 8 | - | Yes | Standard bronchoscope | All patients recovered |
| 12 | Prospective study | Italy | 87 | - | - | Yes | Yes | Disposable bronchoscope | Useful and effective tool |
| 18 | Case report | China | 1 | 1 | 0 | Yes | - | Standard bronchoscope | Optimal PEEP improved bronchoscopy |
| 52 | Case series | South Africa | 4 | 3 | 1 | Yes | - | Standard bronchoscope | Emergency bronchoscopy was safe |
| 65 | Case Report | Japan | 1 | 1 | 0 | Yes | Yes | Fiberoptic and rigid bronchoscope | Safe for patients and HCW |
| 27 | Retrospective study | Belgium | 16 | 11 | 5 | - | Yes | Disposable bronchoscope | Bronchoscopy can be performed safely |
| 62 | Case series | Pakistan | 3 | 2 | 1 | - | Yes | Standard bronchoscope | PDT safe under bronchoscopy |
| 82 | Retrospective study | South Korea | 7 | 6 | 1 | - | Yes | Disposable bronchoscope | Bronchoscopic PDT is safe |
| 79 | Case series | Indonesia | 3 | 2 | 1 | Yes | - | Standard bronchoscope | Helpful in weaning |
| 50 | Multicentre cohort study | Spain | 485 | 357 | 128 | Yes | Yes | 60% Disposable and 40% standard bronchoscopes | Bloody secretions led to death of 496 patients |
| 16 | Interventional study | Italy | 57 | - | - | Yes | - | Flexible bronchoscope | Safe to use bronchoscopy |
| 40 | Case Report | USA | 1 | 0 | 1 | Yes | - | Standard bronchoscope | Patient discharged |
| 36 | Case Report | China | 1 | 1 | 0 | Yes | - | Standard bronchoscope | Patient cured |
| 37 | Case Report | China | 3 | 3 | 0 | Yes | - | Standard bronchoscope | Therapy useful |
| 54 | Case Study | Ireland | 1 | 1 | 0 | Yes | - | Standard bronchoscope | Improved oxygen saturation |
| 72 | Prospective study | USA | 67 | 48 | 19 | - | Yes | Standard bronchoscope | Bronchoscope used to treat complications |
| 56 | Single center retrospective study | Spain | 75 | 54 | 21 | Yes | Yes | Disposable bronchoscope | Bronchoscope is safe invasive technique |
| 24 | Prospective study | Belgium | 59 | - | - | Yes | - | Standard bronchoscope | High-risk bronchoscopic maneuvers are safe |
| 81 | case-series | USA | 12 | 7 | 5 | - | Yes | Disposable bronchoscope | Promoted weaning and safe for HCW |
| 92 | Observational Study | China | 16 | 12 | 4 | Yes | - | Bedside bronchoscopy | Increased in lung O2 and Co2 exchange |
| 30 | Case study | Pakistan | 1 | 1 | 0 | Yes | Yes | Standard bronchoscope | Complete recovery |
| 19 | Case study | China | 1 | 0 | 1 | Yes | - | Standard bronchoscope | Therapeutic bronchoscopy is needed post-LT |
| 66 | Case series | UK | 5 | 5 | 0 | Yes | Yes | Fibreoptic and Rigid bronchoscope | large airway intervention beneficial |
| 22 | Prospective study | India | 61 | 51 | 10 | Yes | Yes | Bedside bronchoscopy | Improved lungs condition after therapeutic suctioning |
| 67 | Retrospective study | USA | 7 | - | - | - | Yes | Standard bronchoscope | Decreased peri & postoperative bleeding |
| 60 | Case series | Portugal | 11 | - | - | Yes | - | Standard bronchoscope | FBB gave a clear therapeutic benefit post CT scan |
| 14 | Retrospective multi-center study | Italy | 109 | 77 | 32 | Yes | Yes | Rigid and flexible Bronchoscopes | Bronchoscopy is a useful technique |
| 80 | Retrospective study | France | 18 | 14 | 4 | Yes | Yes | Disposable bronchoscope | Percutaneous tracheostomy is feasible for physicians |
| 89 | Prospective case series | Indonesia | 3 | 1 | 2 | Yes | - | Disposable bronchoscope | The procedure improved the lung function |
| 83 | Observational study | USA | 18 | 13 | 5 | - | Yes | Disposable bronchoscope | Bronchoscopic PDT safe |
| 13 | Online survey of Global pandemic SARS-CoV-2 bronchoscopy database (GPS-BD) | USA | 289 | 207 | 82 | Yes | Yes | - | Bronchoscopy has reasonable clinical benefit |
| 84 | Prospective single-arm cohort study | USA | 58 | 39 | 19 | - | Yes | Standard bronchoscope | Critically ill patients survived after PDT |
| 78 | Case series | Italy | 2 | 2 | 0 | Yes | - | Standard bronchoscope | Laryngectomies with COVID-19 are complex |
| 53 | Case series | India | 1 | 1 | 0 | Yes | - | Rigid bronchoscopy | Patient discharged |
| 38 | Multicenter retrospective study | Italy | 43 | 31 | 12 | Yes | - | Standard bronchoscope | Bronchoscopic BAL isolation effective method |
| 39 | Case Series | Greece | 1 | 1 | 0 | Yes | - | Standard bronchoscope | Transferred from ICU |
| 26 | Retrospective study | Italy | 21 | 17 | 4 | Yes | - | Standard bronchoscope | Surfactant delivery through bronchoscopy is effective therapy |
| 29 | Retrospective case series | USA | 3 | 2 | 1 | Yes | Yes | Standard bronchoscope | Feasible to perform PDT on intubated patient with optimal PEEP and FiO2 |
| 42 | Case report | USA | 1 | 1 | 0 | Yes | - | Standard bronchoscope | Successfully managed using serial therapeutic bronchoscopy |
| 15 | Retrospective cohort study | USA | 37 | 22 | 15 | Yes | Yes | Disposable bronchoscope | Benefit to patients with PDT placement |
| 68 | Proof of concept study | USA | 11 | 7 | 4 | - | Yes | Disposable bronchoscope | Efficacious and adaptable protocol for bronchoscopy guided PDT |
| 76 | Case report | India | 1 | 1 | 0 | Yes | - | Standard bronchoscope | Complete resolution of lung failure |
| 87 | Retrospective study | USA | 12 | 9 | 3 | - | Yes | Fibreoptic bronchoscope | Modified PDT are safe in patients |
| 74 | Case report | Japan | 1 | 1 | 0 | Yes | - | Standard bronchoscope | Successful treatment patient discharged |
| 85 | Case study | USA | 1 | 1 | 0 | - | Yes | Flexible bronchoscope | Patient has sufficiently recovered |
| 45 | Observational study | South Africa | 14 | 9 | 5 | Yes | - | Flexible fibreoptic bronchoscopy | Oxygenation improved |
| 71 | Retrospective observational study | Italy | 120 | 80 | 40 | - | Yes | Video-assisted fibro-bronchoscopy | Procedure was safe for all HCW |
| 28 | Observational study | Spain | 93 | - | - | Yes | - | Disposable bronchoscopy | Patients can be benefit from bronchoscopy |
| 59 | Case series | USA | 8 | 6 | 2 | Yes | - | Disposable bronchoscope | Potentially safe for patient care |
| 88 | Prospective study | UK | 38 | 31 | 7 | - | Yes | Disposable bronchoscope | All procedures were successful |
| 63 | Retrospective study | USA | 3 | 2 | 1 | - | Yes | Standard bronchoscope | Modified percutaneous tracheostomy is feasible |
| 43 | Case series | Indonesia | 3 | 3 | 0 | Yes | - | Standard bronchoscope | Significant clinical improvement seen |
| 44 | Case report | Indonesia | 1 | 1 | 0 | Yes | - | Standard bronchoscope | The patient survived |
| 41 | Case series | USA | 11 | 9 | 2 | Yes | - | Standard bronchoscope | Patient rescued from ETT block |
| 11 | A prospective randomized controlled trial | China | 58 | 33 | 25 | - | Yes | Portable bronchoscope | HFNO provided a shorter intubation for critically ill. |
| 64 | Single center prospective study | UK | 72 | 51 | 21 | - | Yes | Standard bronchoscope | OT and PDT both are equally favorable. |
| 69 | Retrospective study | USA | 27 | 23 | 4 | Yes | - | Standard bronchoscope | Judge well before using ECMO. |
|  |  | Majority of studies from USA 23, Italy 10, China 9. | 2558 | 1563 male | 598 female |  |  | DB 17 studies, RB 1 studies, SB 49 studies, 1 not reported, 3 studies RB+SB, and 1 study used SB+DB | Safety 17 studies, 5 studies no comments on bronchoscopy, 50 studies on efficacy |

